# Changes in hypothalamic subunits volume and their association with metabolic parameters and gastrointestinal appetite-regulating hormones following bariatric surgery

**DOI:** 10.1101/2024.08.30.24312638

**Authors:** Amélie Lachance, Justine Daoust, Mélissa Pelletier, Alexandre Caron, André C. Carpentier, Laurent Biertho, Josefina Maranzano, André Tchernof, Mahsa Dadar, Andréanne Michaud

## Abstract

**Background:** Some nuclei of the hypothalamus are known for their important roles in maintaining energy homeostasis and regulating food intake. Moreover, obesity has been associated with hypothalamic inflammation and morphological alterations, as indicated by increased volume. However, the reversibility of these changes after bariatric surgery-induced weight loss remains underexplored.

**Objective:** The aim of this study was to characterize volume changes in hypothalamic subunits up to two years following bariatric surgery and to determine whether these differences were associated with changes in metabolic parameters and levels of gastrointestinal appetite-regulating hormone levels.

**Methods:** Participants with severe obesity undergoing bariatric surgery were recruited. They completed high-resolution T1-weighted brain magnetic resonance imaging (MRI) before bariatric surgery and at 4, 12 and 24 months post-surgery. Blood samples collected during the fasting and postprandial states were analyzed for glucagon-like peptide 1 (GLP-1), peptide YY (PYY), and ghrelin concentrations. The hypothalamus was segmented into 5 subunits per hemisphere using a publicly available automated tool. Linear mixed-effects models were employed to examine volume changes between visits and their associations with variables of interest.

**Results:** A total of 73 participants (mean age 44.5 ± 9.1 years, mean BMI 43.5 ± 4.1 kg/m^2^) were included at baseline. Significant volume reductions were observed in the whole left hypothalamus 24 months post-surgery. More specifically, decreases were noted in both the left anterior-superior and left posterior subunits at 12 and 24 months post-surgery (all p<0.05, after FDR correction). These reductions were significantly associated with the percentage of total weight loss (both subunits p<0.001), improvements in systolic blood pressure (both subunits p<0.05), and an increase in postprandial PYY (both subunits p<0.05).

**Conclusion:** These results suggest that some hypothalamic morphological alterations observed in the context of obesity could potentially be reversed with bariatric surgery induced-weight loss.

## 1 Introduction

Food intake involves a complex, simultaneous interplay of various brain regions (Dagher, 2012). Despite its modest 1% contribution to total brain weight on average, the hypothalamus plays a central role in regulating this process (Dagher, 2012; Clifford B. Saper, 2012). Acting as a control center, it integrates signals from a myriad of afferent nerves within the body and coordinates their transmission to cortical and subcortical grey matter (C. B. Saper & Lowell, 2014). Therefore, the hypothalamus contributes to several critical biological mechanisms essential for basic survival functions, including energy metabolism and the homeostatic control of food intake (C. B. Saper & Lowell, 2014). These functions are orchestrated by a dozen specific nuclei within the hypothalamus, which are characterized by distinct cellular populations (Bedont, Newman, & Blackshaw, 2015; Billot et al., 2020; Xie & Dorsky, 2017). Among these are the arcuate, lateral, paraventricular, dorsomedial and ventromedial nuclei, all implicated in the regulation of food intake (Caron & Richard, 2017).

Given its fundamental role in metabolic homeostasis, the hypothalamus has been extensively studied to elucidate potential disruptions associated with obesity. Most of these studies have been conducted in animal models, with relatively limited investigations conducted in humans. Challenges associated with the small size of the hypothalamus have limited the use of magnetic resonance imaging (MRI) techniques until recent advancements. Manual delineation of its boundaries, which requires expertise and a valuable amount of time (Schindler et al., 2013), has now been supplanted by the development of automated segmentation techniques, facilitating the study of larger cohorts (Billot et al., 2020; Rodrigues et al., 2022). The automated segmentation tool developed by Billot *et al*. divides the hypothalamus into five subunits per hemisphere, offering nuanced insights into its functioning (Billot et al., 2020). Using this tool, two recent studies showed morphological alterations in the hypothalamus among individuals living with overweight or obesity, with increased volume for the whole hypothalamus and for specific subunits (Alzaid et al., 2024; Brown, Westwater, Seidlitz, Ziauddeen, & Fletcher, 2023). However, the mechanisms underlying these findings and whether these hypothalamic alterations in individuals with obesity are permanent or reversible following interventions targeting weight loss and cardiometabolic improvements remain unexplored.

Bariatric surgery, known for inducing significant and sustained weight loss alongside metabolic improvements, presents a unique opportunity to study long-term effects on brain integrity in a prospective setting (Maciejewski et al., 2016; O’Brien et al., 2019; Salminen et al., 2022). Significant changes in both white and grey matter densities have been documented post-bariatric surgery (Michaud et al., 2020; Rullmann et al., 2018; Tuulari et al., 2016; Wang et al., 2020). Previous studies have also examined the impact of bariatric surgery on hypothalamic morphological changes using voxel-based morphometry (VBM), with only one reporting volume changes in specific subregions (Rullmann et al., 2019; van de Sande-Lee et al., 2020). A recent study employing an automated segmentation tool, which is considered more reliable than VBM techniques for assessing volume in such a small region of the brain (Arkink et al., 2017), demonstrated reductions in hypothalamic volume one year post-surgery, particularly in different subunits of the left hypothalamus (Pané et al., 2024). However, a study with longer-term follow-up is needed to explore the sustainability of the effects of weight-loss and concomitant cardiometabolic improvements on hypothalamic morphometric changes.

The mechanisms underlying hypothalamic morphological alterations in the context of obesity remain unclear. Various hypotheses have been explored over the years, including hypothalamic inflammation and gliosis (Schur et al., 2015; Sewaybricker, Huang, Chandrasekaran, Melhorn, & Schur, 2023; Thaler, Guyenet, Dorfman, Wisse, & Schwartz, 2013) as well as a decreased level of brain vascularization and cerebral hypoperfusion (Alfaro et al., 2018; Bettcher et al., 2013; Willeumier, Taylor, & Amen, 2011). Some gastrointestinal hormones could also play a role in the remodeling process of the hypothalamus, as they exert their effects by binding to receptors within the hypothalamus (Al Massadi, López, Tschöp, Diéguez, & Nogueiras, 2017; Farr et al., 2016; Jensen et al., 2018; Karra, Chandarana, & Batterham, 2009; Li et al., 2023) or by acting on neurons projecting into hypothalamic nuclei (Biddinger, Lazarenko, Scott, & Simerly, 2020). Moreover, concomitant changes in hunger and satiety hormones, such as ghrelin, glucagon-like peptide 1 (GLP-1), and peptide YY (PYY), have been reported in obesity (Aukan, Coutinho, Pedersen, Simpson, & Martins, 2023) and following bariatric surgery (Huang et al., 2023). It remains unknown how changes in peripheral gut hormones are associated with hypothalamic morphometric changes following bariatric surgery.

In this prospective study, we aimed to characterize changes in hypothalamic volume and its subunits up to two years following bariatric surgery by employing an automated segmentation tool (Billot et al., 2020). As a secondary objective, we examined whether these changes were associated with weight loss, metabolic parameters, and variations in gastrointestinal appetite-regulating hormone levels, including ghrelin, GLP-1, and PYY. We hypothesized that bariatric surgery leads to reductions in total hypothalamus volume and specific subunits, and these changes are associated with the magnitude of weight loss, as well as improvements in cardiometabolic parameters and gastrointestinal appetite-regulating hormone levels.

## 2 Methods

### 2.1 Participants

Ninety-two participants scheduled to undergo bariatric surgery at the *Institut universitaire de cardiologie et de pneumologie de Québec-Université Laval* (IUCPQ-UL) in Québec, Canada, were recruited (Figure S1). These participants were part of a larger prospective study, which aimed at investigating the determinants of metabolic recovery following bariatric surgeries. The protocol has been previously described in detail (Michaud et al., 2020). Briefly, participants had to be 18 to 60 years old and meet the NIH Guidelines for bariatric surgery ("Gastrointestinal surgery for severe obesity: National Institutes of Health Consensus Development Conference Statement," 1992). Exclusion criteria included gastrointestinal diseases, such as irritable bowel syndrome or gastro-intestinal ulcers, cirrhosis or albumin deficiency, neurological illnesses, uncontrolled high blood pressure, use of any medication that can affect the central nervous system, previous bariatric or brain surgery, severe food allergy, current pregnancy and substance or alcohol abuse. Participants with any contraindications for MRI, including claustrophobia, metal embedded in the body, or the presence of an implanted medical device, were also excluded. The protocol was approved by the Research Ethics Committee of the *Centre de recherche de l’IUCPQ-UL* (no. 2016-2569, 21237). A written consent form was signed by each participant at their first visit.

### 2.2 Surgical procedures

Participants received either a sleeve gastrectomy (SG), a Roux-en-Y gastric bypass (RYGB) or a biliopancreatic diversion with duodenal switch (BPD-DS). All surgeries were performed laparoscopically. Regarding SG, a 250 cm^3^ vertical gastrectomy was performed with a 34-44 French Bougie starting 7-8 cm from the pylorus to the His angle (Biertho et al., 2016). For the RYGB, a 30-50 cm^3^ proximal gastric pouch was first created and then connected to the proximal small intestine by bypassing the first 100 cm (Zeighami et al., 2021). Finally, the BPD-DS was performed by first doing a 250 cm^3^ SG and thereafter, the duodenum was transected about 4 cm distal from the pylorus and anastomosed to a 250-cm alimentary limb, with a 100-cm common channel (Biertho et al., 2010).

### 2.3 Study design and experimental procedures

Outcomes were assessed prior to the surgery, and at 4, 12, and 24 months post-op. An identical protocol was followed at each visit. Participants were asked to fast for 12 hours before the visit. Weight and body composition were measured by bioimpedance (InBody520, body composition analyzer, Biospace, Los Angeles, California or Tanita DC-430U, Arlington Heights, IL). Anthropometric measures (waist, hip, and neck circumferences) and blood pressure [systolic (SBP) and diastolic (DBP)] were measured using standardized procedures (Michaud et al., 2020). The percentage of total weight loss (%TWL; [(baseline weight - follow-up weight)/baseline weight] x 100%) and the percentage of excess weight loss (%EWL; [(baseline weight - follow-up weight)/(pre-surgery weight − ideal weight for a body mass index (BMI) of 23 kg/m^2^)] x 100%) were calculated.

### 2.4. Plasma lipid profile, glucose homeostasis, and gastrointestinal appetite-regulating hormones

Blood samples were taken after a 12-hour overnight fast and approximately one hour after the consumption of a standard nutritional beverage [237 ml Boost Original, Nestlé Health Science (240 kcal, 41g of carbohydrates, 10g of proteins and 4g of lipids)]. Blood samples were collected into chilled tubes with appropriate enzymatic inhibitors (dipeptidyl peptidase-IV inhibitor, aprotinin, or HCl) (Millipore Sigma Canada, Ontario). All samples were immediately kept at 4°C before being centrifuged, and then stored at -80°C. Fasted acylated and non-acylated ghrelin were measured using ELISA kits (Bertin Pharma; A05106 and A05119, respectively) and summed to obtain total ghrelin. Levels of post-prandial GLP-1 and postprandial PYY were measured by multiplex assay (Millipore Sigma Canada, Ontario; HMH3-34K) (Richard et al. 2022). Plasma levels of cholesterol, high-density lipoproteins (HDL), low-density lipoproteins (LDL), triglycerides, glucose and insulin were measured at each visit as part of the routine monitoring of patients. The homeostatic model assessment for insulin resistance (HOMA-IR) was calculated [fasting glucose (mmol/L) x fasting insulin (pmol/L) / 135) (Matthews et al., 1985).

### 2.5 MRI acquisition

T1-weighted three-dimensional (3D) turbo echo images of the brain were acquired using a 3T whole-body MRI scanner (Philips, Ingenia, Philips Medical Systems) equipped with a 32-channel head coil at the *Centre de recherche de l’IUCPQ-UL*. The following parameters were used: 176 sagittal 1.0 mm slices, TR/TE = 8.1/3.7 ms, field of view (FOV) = 240 x 240 mm^2^, and voxel size = 1 x 1 x 1 mm.

### 2.6 MRI data analysis and hypothalamus segmentation

Standard preprocessing steps, including image denoising (Coupe et al., 2008), intensity non uniformity correction (Sled, Zijdenbos, & Evans, 1998) and image intensity normalization into range (0–100) using histogram matching, were first applied. Images were then linearly (using a nine-parameter rigid registration) registered to an average brain template (MNI ICBM152-2009c) using MNI MINC tools (http://www.bic.mni.mcgill.ca/ServicesSoftware/MINC). This step maps all the participants’ brain onto the same standard space, improving the performance of the downstream segmentation pipeline and eliminating the need to adjust for total intracranial volume. All registrations were visually controlled to ensure their accuracy. Segmentation of the hypothalamus was performed on these standardized images using an open source method based on a deep convolutional neural network (Billot et al., 2020). This automated segmentation tool has been previously validated and used to perform hypothalamus segmentation in similar applications (Alzaid et al., 2024; Brown et al., 2023; Pané et al., 2024; Ruzok et al., 2022), and is part of the FreeSurfer brain segmentation software. This tool allows the segmentation of the hypothalamus in 5 subunits per hemisphere: anterior-inferior, anterior-superior, posterior, tubular inferior and tubular superior (Figure 1). Each of these subunits includes two to five hypothalamic nuclei. Using FSLeyes, a tool from the FMRIB Software Library (Jenkinson, Beckmann, Behrens, Woolrich, & Smith, 2012), every individual segmentation (n=245) was visually inspected for quality control based on a protocol published by Bochetta *et al*. (Bocchetta et al., 2015). Segmentations with irregular shapes (n=56), such as holes and fragmented subunits, were considered low quality and were excluded from further analyses (Figure S1). Additionally, we created a segmentation quality variable that we included as a covariate in our statistical models to further assess the robustness of our results. Images showing overall good segmentation but with minor defects, such as some missing a few voxels lining the 3rd ventricle, were classified as moderate-quality segmentations (n=64) but were still included in the analyses. The remaining segmentations were classified as high-quality (n=125).

**Figure 1.**
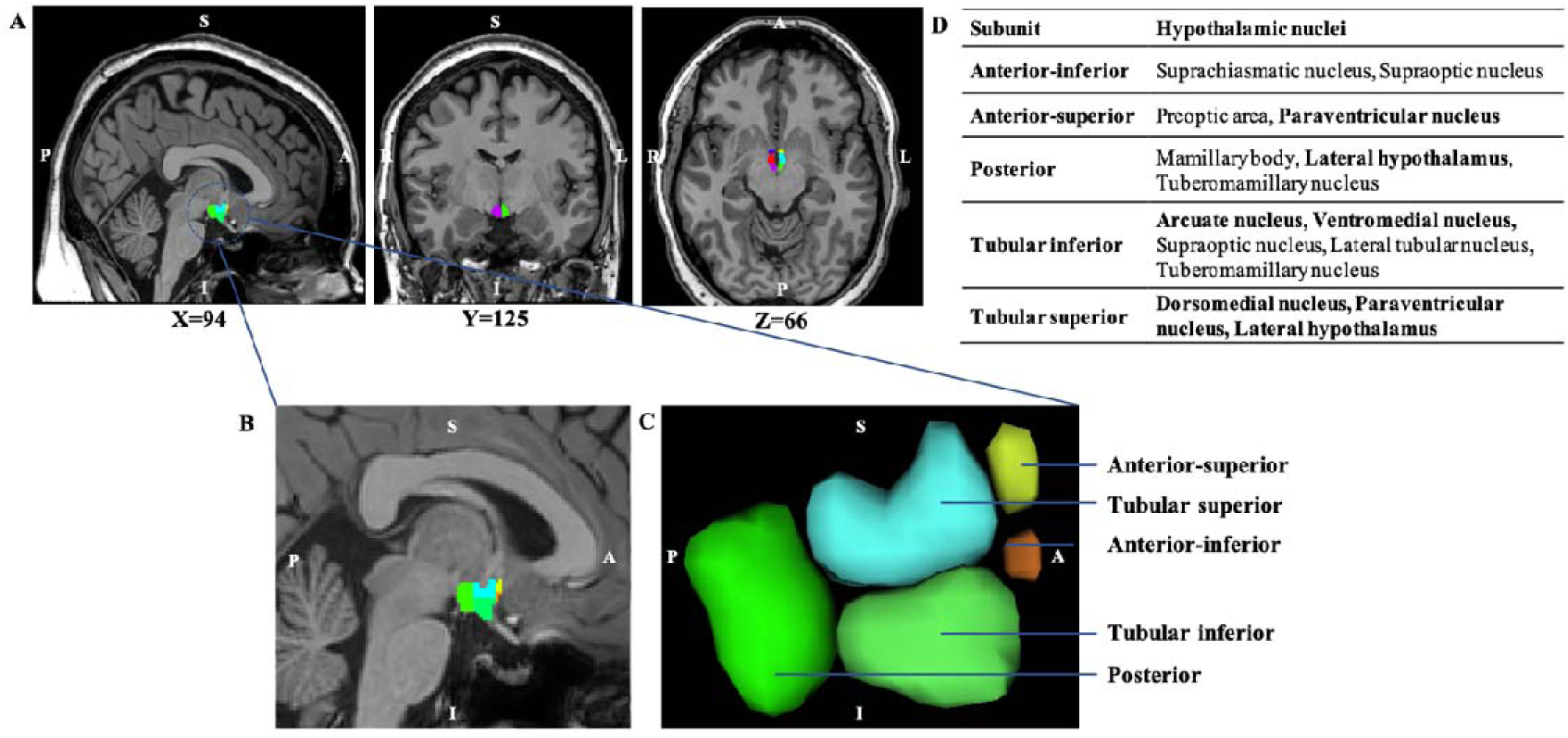
Visual representation of the hypothalamic segmentation. A) Sagittal, Coronal and Axial brain slices showing hypothalamic segmentation and different hypothalamic subunits. B) Expanded view of the sagittal plane showing the left hypothalamus. C) Three-dimensional view of the left hypothalamus. D) Tables presenting the five hypothalamic subunits, and the corresponding hypothalamic nuclei included within them. Principal nuclei involved in food intake are highlighted in bold. ‘L’ denotes left, and ‘R’ denotes right. ‘P’ denotes posterior, and ‘A’ denotes anterior.

### 2.7 Statistical analyses

Participants’ demographic data, cardiometabolic variables, and changes in gastrointestinal appetite-regulating hormones following bariatric surgery were assessed using Chi-squared tests for categorical variables and linear mixed-effects models for continuous variables. Linear mixed-effects models (Model I) were performed to examine hypothalamic volume changes following bariatric surgery. Participants were identified as categorical random variables, and age, sex, initial BMI, surgery type and session were added as fixed effects.

**Model I:** Hypothalamic volume ∼ Session + Age + Sex + BMI_baseline_ + Surgery type + (1| Participant)

Linear mixed-effects models (Model II) were used to examine the associations between the changes in cardiometabolic variables (total weight loss, percentage of fat mass, SBP, HOMA-IR, plasmatic levels of triglycerides and HDL-C), and total left and right hypothalamus volume and their subunits. Participants were added to the model as categorical random variables, and age, sex, initial BMI and surgery type as fixed effects.

**Model II:** Hypothalamic volume ∼ cardiometabolic variable + Age + Sex + BMI_baseline_ + Surgery type + (1| Participant)

Linear mixed-effects models (Model III) were also used to examine the associations between the changes in gastrointestinal appetite-regulating hormones (fasting total ghrelin and acylated ghrelin, as well as postprandial GLP-1 and PYY) and total left or right hypothalamus volume and their subunits. Participants were added to the model as categorical random variables, and age, sex, initial BMI and surgery type as fixed effects.

**Model III:** Hypothalamic volume ∼ gastrointestinal appetite-regulating hormones + Age + Sex + BMI_baseline_ + Surgery type + (1| Participant)

The variables referred to as “cardiometabolic variable” or “gastrointestinal appetite-regulating hormones” in Models II and III represent the prospective measurements of the absolute levels of each variable, collected and entered for all time points. Due to the limited sample size and multicollinearity, the effect of each cardiometabolic or hormonal variable was examined separately.

Additional analyses were conducted to determine if segmentation quality significantly impacted the results. First, volumes of high-quality segmentations were compared to volumes of moderate-quality segmentations for each visit separately. T-tests were used for normally distributed variables with equal variance, whereas Welch’s tests were used for unequal variance. Variables not following a normal distribution were first transformed (log or BoxCox transformation). If a normal distribution could not be achieved after transformations, a non-parametric test (Wilcoxon test) was used. Furthermore, the segmentation quality variable was integrated as a fixed effect covariate in Model I to determine if segmentation quality (high versus moderate) significantly impacted the results.

All results were corrected for multiple comparisons with the False Discovery Rate (FDR) controlling approach (p < 0.05) using the Benjamini-Hochberg procedure. Statistical analyses were performed using JMP Pro version 16.0 (SAS Institute Inc., Cary, NC, USA).

## 3 Results

### 3.1 Clinical characteristics of the participants

Table 1 shows the characteristics of the participants. After quality control of the hypothalamus segmentations, a total of 73 participants (mean age 44.5 ± 9.1 years, mean BMI 43.5 ± 4.1 kg/m^2^) were included at baseline (Figure S1). The study population consisted mainly of females (71%). SG was the most frequently performed procedure. As expected, participants had significant weight loss following bariatric surgery (p < 0.0001). They also showed improvements in blood pressure (systolic and diastolic, both p < 0.0001), improvements in glucose homeostasis markers (fasting glycemia, fasting insulin, and HOMA-IR index, all p < 0.0001) and lipid profile (total cholesterol, p = 0.009; HDL-C, p < 0.0001; and triglycerides, p < 0.0001). Total and acylated fasting ghrelin levels were significantly decreased post-surgery (both p < 0.0001), while postprandial GLP-1 and PYY levels were significantly increased (p = 0.029 and p < 0.0001, respectively).

**Table 1.**
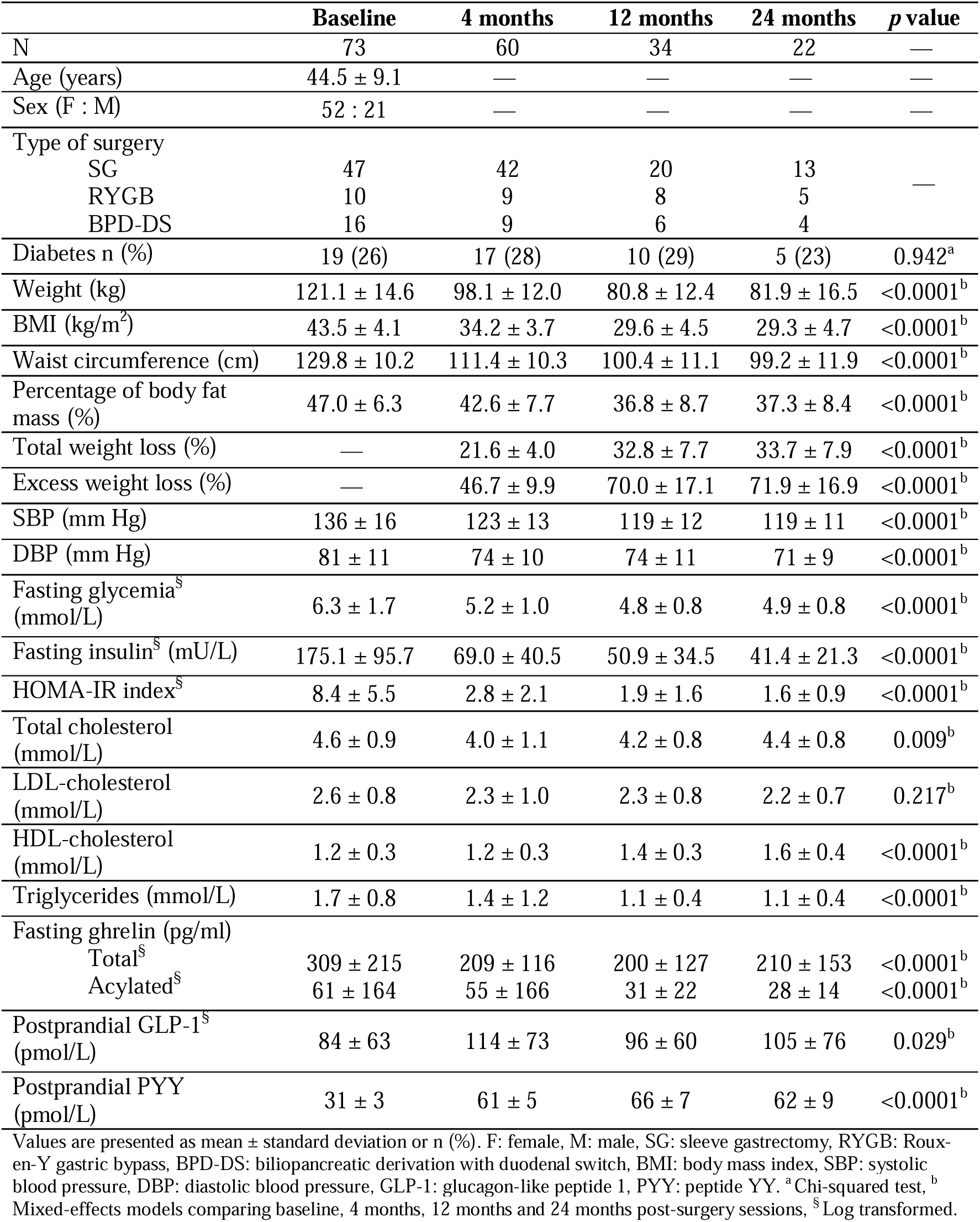
Characteristics of participants.

|  | Baseline | 4 months | 12 months | 24 months | p value |
| --- | --- | --- | --- | --- | --- |
| N | 73 | 60 | 34 | 22 | — |
| Age (years) | 44.5 ± 9.1 | — | — | — | — |
| Sex (F : M) | 52 : 21 | — | — | — | — |
| Type of surgery |  |  |  |  |  |
| SG | 47 | 42 | 20 | 13 | — |
| RYGB | 10 | 9 | 8 | 5 |  |
| BPD-DS | 16 | 9 | 6 | 4 |  |
| Diabetes n (%) | 19 (26) | 17 (28) | 10 (29) | 5 (23) | 0.942<sup>a</sup> |
| Weight (kg) | 121.1 ± 14.6 | 98.1 ± 12.0 | 80.8 ± 12.4 | 81.9 ± 16.5 | <0.0001<sup>b</sup> |
| BMI (kg/m<sup>2</sup>) | 43.5 ± 4.1 | 34.2 ± 3.7 | 29.6 ± 4.5 | 29.3 ± 4.7 | <0.0001<sup>b</sup> |
| Waist circumference (cm) | 129.8 ± 10.2 | 111.4 ± 10.3 | 100.4 ± 11.1 | 99.2 ± 11.9 | <0.0001<sup>b</sup> |
| Percentage of body fat mass (%) | 47.0 ± 6.3 | 42.6 ± 7.7 | 36.8 ± 8.7 | 37.3 ± 8.4 | <0.0001<sup>b</sup> |
| Total weight loss (%) | — | 21.6 ± 4.0 | 32.8 ± 7.7 | 33.7 ± 7.9 | <0.0001<sup>b</sup> |
| Excess weight loss (%) | — | 46.7 ± 9.9 | 70.0 ± 17.1 | 71.9 ± 16.9 | <0.0001<sup>b</sup> |
| SBP (mm Hg) | 136 ± 16 | 123 ± 13 | 119 ± 12 | 119 ± 11 | <0.0001<sup>b</sup> |
| DBP (mm Hg) | 81 ± 11 | 74 ± 10 | 74 ± 11 | 71 ± 9 | <0.0001<sup>b</sup> |
| Fasting glycemia<sup>§</sup> (mmol/L) | 6.3 ± 1.7 | 5.2 ± 1.0 | 4.8 ± 0.8 | 4.9 ± 0.8 | <0.0001<sup>b</sup> |
| Fasting insulin<sup>§</sup> (mU/L) | 175.1 ± 95.7 | 69.0 ± 40.5 | 50.9 ± 34.5 | 41.4 ± 21.3 | <0.0001<sup>b</sup> |
| HOMA-IR index<sup>§</sup> | 8.4 ± 5.5 | 2.8 ± 2.1 | 1.9 ± 1.6 | 1.6 ± 0.9 | <0.0001<sup>b</sup> |
| Total cholesterol (mmol/L) | 4.6 ± 0.9 | 4.0 ± 1.1 | 4.2 ± 0.8 | 4.4 ± 0.8 | 0.009<sup>b</sup> |
| LDL-cholesterol (mmol/L) | 2.6 ± 0.8 | 2.3 ± 1.0 | 2.3 ± 0.8 | 2.2 ± 0.7 | 0.217<sup>b</sup> |
| HDL-cholesterol (mmol/L) | 1.2 ± 0.3 | 1.2 ± 0.3 | 1.4 ± 0.3 | 1.6 ± 0.4 | <0.0001<sup>b</sup> |
| Triglycerides (mmol/L) | 1.7 ± 0.8 | 1.4 ± 1.2 | 1.1 ± 0.4 | 1.1 ± 0.4 | <0.0001<sup>b</sup> |
| Fasting ghrelin (pg/ml) |  |  |  |  |  |
| Total<sup>§</sup> | 309 ± 215 | 209 ± 116 | 200 ± 127 | 210 ± 153 | <0.0001<sup>b</sup> |
| Acylated<sup>§</sup> | 61 ± 164 | 55 ± 166 | 31 ± 22 | 28 ± 14 | <0.0001<sup>b</sup> |
| Postprandial GLP-1<sup>§</sup> (pmol/L) | 84 ± 63 | 114 ± 73 | 96 ± 60 | 105 ± 76 | 0.029<sup>b</sup> |
| Postprandial PYY (pmol/L) | 31 ± 3 | 61 ± 5 | 66 ± 7 | 62 ± 9 | <0.0001<sup>b</sup> |

Values are presented as mean ± standard deviation or n (%). F: female, M: male, SG: sleeve gastrectomy, RYGB: Roux-en-Y gastric bypass, BPD-DS: biliopancreatic derivation with duodenal switch, BMI: body mass index, SBP: systolic blood pressure, DBP: diastolic blood pressure, GLP-1: glucagon-like peptide 1, PYY: peptide YY. <sup>a</sup>Chi-squared test, <sup>b</sup>Mixed-effects models comparing baseline, 4 months, 12 months and 24 months post-surgery sessions, <sup>§</sup>Log transformed.

No significant differences in age, sex, and surgery type were observed between the high-quality segmentation group and the moderate-quality segmentation group for all sessions (Table S1). However, participants in the moderate-quality segmentation group had a higher BMI at baseline and a lower total weight loss at 12 months post-surgery compared to participants in the high-quality segmentation group (p=0.0307 and p=0.0226, respectively; Table S1). A higher percentage of participants with Type 2 diabetes was observed in the high-quality segmentation group compared to the moderate-quality group at 4 months post-surgery (p=0.0027, Table S1).

### 3.2 Hypothalamic volume changes following bariatric surgery

Figure 2a shows the T-values from the mixed effects-models (Model I), contrasting the follow-up regional volumes against their baseline values. A significant decrease in the whole volume of the left hypothalamus was observed 24 months post-surgery compared to baseline (p = 0.0007, Figure 2b). More specifically, a local decrease in volume was observed in the left anterior-superior subunit 4 months post-surgery compared to baseline (p < 0.05 after FDR correction; Figure 2c). This decrease in volume was maintained at 12 months and 24 months post-surgery (p < 0.05 and p < 0.01 after FDR correction; Figure 2c). Additionally, a significant decrease in the volume of the left posterior subunit was also observed 12 and 24 months post-surgery compared to baseline (both p < 0.001 after FDR correction, respectively; Figure 2d). No significant changes in volume were found in the right hypothalamus and its subunits following bariatric surgery. When comparing the volumes of the left and right hypothalamus and their subunits of the high- and moderate-quality groups at each visit, the only significant difference between the two groups was for the whole left hypothalamus (Table S2). Participants in the moderate-quality segmentation group had, on average, a higher volume than those in the high-quality segmentation group. No significant difference for the other subunits was observed after FDR correction. Furthermore, adding the segmentation quality variable as a fixed effect in Model I did not change the results regarding volume changes for the left and right hypothalamus and their subunits (Table S3).

**Figure 2.**
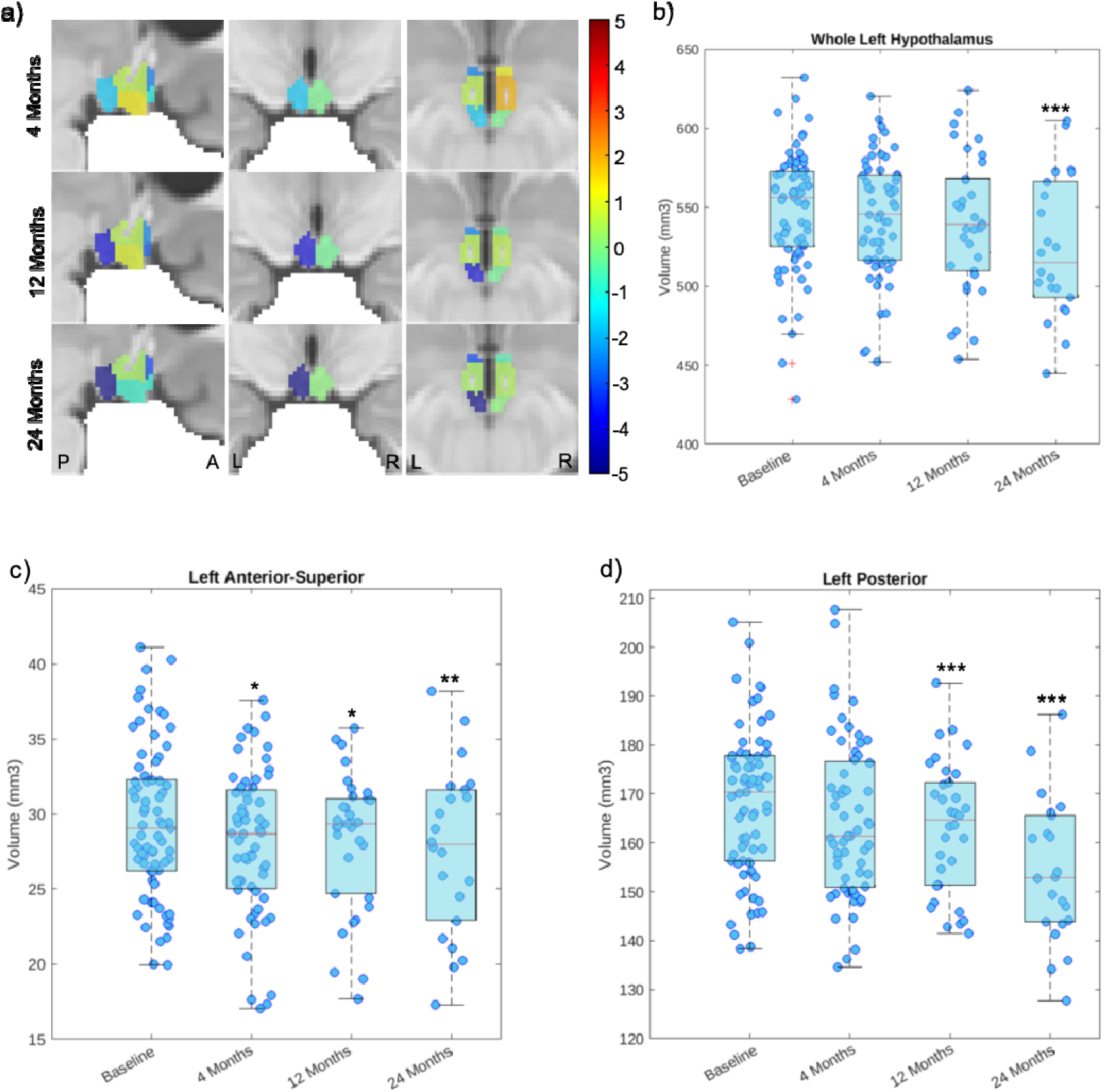
Changes in hypothalamus subunits 4, 12 and 24 months following bariatric surgery compared to baseline. Panel a) shows the T-values from the mixed-effects models, contrasting the follow-up regional volumes against their baseline values. Warmer colors (e.g. orange and red) indicate an increase after surgery, while colder colors (e.g. blue and purple) indicate a decrease after surgery. ‘L’ denotes left, and ‘R’ denotes right. ‘P’ denotes posterior, and ‘A’ denotes anterior. Significant changes in volumes are observed in b) the whole left hypothalamus, c) the left anterior-superior subunit and d) the left posterior subunit. * p < 0.05, ** p < 0.01, *** p < 0.001 versus baseline.

### 3.3 Associations between changes in hypothalamic volumes and cardiometabolic variables following bariatric surgery

We used a mixed-effects model (Model II) to examine the association between changes in hypothalamic volumes and cardiometabolic variables. Significant associations were found between the volume of the whole left hypothalamus and the percentage of total weight loss (□ = -39.88, p = 0.0008), as well as the percentage of fat mass (□ = 1.06, p = 0.0021), wherein a more pronounced decrease in hypothalamic volume was associated with greater total weight loss and a reduced percentage of fat mass following bariatric surgery (Table 2). Associations between the percentage of total weight loss and specific subunits were also observed, including the left anterior-inferior (□ = -3.83, p = 0.0223, after FDR correction), left anterior-superior (□ = -5.47, p = 0.0002, after FDR correction), and left posterior subunits (□ = -20.08, p < 0.0001, after FDR correction). Conversely, no significant association was observed between changes in the volume of the right hypothalamus and weight loss.

**Table 2.**
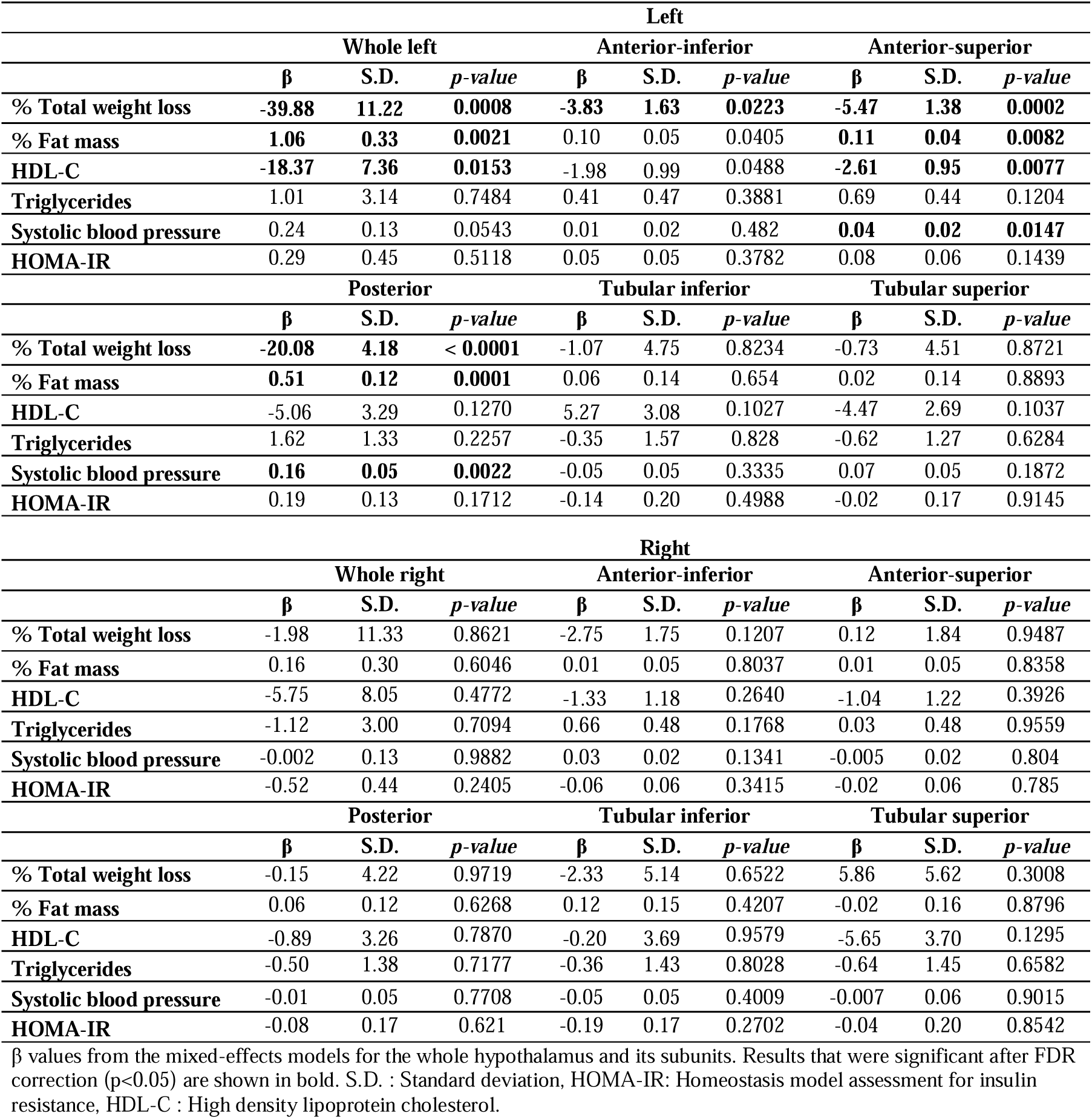
Associations between changes in hypothalamic volumes and metabolic parameters following bariatric surgery.

Reduced volumes in the whole left hypothalamus and in the left anterior-superior subunit were associated with increased levels of HDL-C (□ = -18,37, p = 0.0153 and □ = -2.61, p = 0.0077, respectively, after FDR correction; Table 2), suggesting that improvement in HDL-C is associated with reduced volumes. No significant associations were found between hypothalamic volumes and other lipid profile markers, including triglycerides, LDL-C and total cholesterol (Table 2, results for LDL-C and total cholesterol not shown). Decreased volumes in the left anterior-superior and left posterior subunits were associated with reductions in systolic blood pressure (□ = 0.04, p = 0.0147 and □ = 0.16, p = 0.0022, respectively, after FDR correction; Table 2). No significant associations were found between change in hypothalamic volumes and indicators of glucose homeostasis, namely HOMA-IR, fasting glycemia and fasting insulin (Table 2, results for glycemia and insulin not shown).

### 3.4 Associations between hypothalamic volumes and gastrointestinal appetitive hormone levels following bariatric surgery

A decreased volume in the left anterior-superior and in the left posterior subunits following bariatric surgery was associated with an increase in postprandial PYY (□ = -0.03, p = 0.0034 and □ = -0.10, p = 0.0001, respectively, after FDR correction; Table 3). Only the volume change in the right anterior-inferior subunit showed a positive association with changes in total ghrelin levels (□ = 1.71, p = 0.0055, after FDR correction; Table 3), whereas no association was found with the acylated form of ghrelin. Reduced volumes in the whole right hypothalamus and in the right anterior-inferior subunit were negatively associated with changes in levels of postprandial GLP-1 (□ = -0.09, p = 0.0057 and □ = -0.01, p = 0.0076, respectively, after FDR correction; Table 3).

**Table 3.** Associations between hypothalamic volumes and variations in gastrointestinal appetite-regulating hormone levels following bariatric surgery.

| Left |  |  |  |  |  |  |  |  |  |
| --- | --- | --- | --- | --- | --- | --- | --- | --- | --- |
|  | Whole left |  |  | Anterior-inferior |  |  | Anterior-superior |  |  |
| | $\beta$ | S.D. | *p-value* | $\beta$ | S.D. | *p-value* | $\beta$ | S.D. | *p-value* |
| Ghrelin, total | 5.28 | 4.41 | 0.2337 | 0.39 | 0.56 | 0.4907 | 0.33 | 0.55 | 0.5516 |
| Ghrelin, acylated | 3.97 | 4.75 | 0.4043 | 0.12 | 0.56 | 0.8303 | 0.46 | 0.61 | 0.453 |
| GLP-1 | -0.04 | 0.03 | 0.2490 | 0.005 | 0.005 | 0.3266 | -0.002 | 0.005 | 0.6376 |
| PYY | -0.11 | 0.07 | 0.0918 | 0.007 | 0.009 | 0.4346 | **-0.03** | **0.009** | **0.0034** |
|  | Posterior |  |  | Tubular inferior |  |  | Tubular superior |  |  |
| | $\beta$ | S.D. | *p-value* | $\beta$ | S.D. | *p-value* | $\beta$ | S.D. | *p-value* |
| Ghrelin, total | 1.50 | 1.79 | 0.4027 | 1.45 | 1.92 | 0.4545 | -1.04 | 1.77 | 0.5567 |
| Ghrelin, acylated | -0.57 | 1.92 | 0.7654 | 3.91 | 2.00 | 0.0538 | -0.34 | 1.76 | 0.8488 |
| GLP-1 | -0.04 | 0.01 | 0.0119 | -0.005 | 0.01 | 0.7087 | -0.01 | 0.01 | 0.3292 |
| PYY | **-0.10** | **0.02** | **0.0001** | 0.01 | 0.03 | 0.6260 | -0.03 | 0.03 | 0.2780 |
| Right |  |  |  |  |  |  |  |  |  |
|  | Whole right |  |  | Anterior-inferior |  |  | Anterior-superior |  |  |
| | $\beta$ | S.D. | *p-value* | $\beta$ | S.D. | *p-value* | $\beta$ | S.D. | *p-value* |
| Ghrelin, total | 1.44 | 4.5 | 0.75 | **1.71** | **0.61** | **0.0055** | -0.02 | 0.60 | 0.9796 |
| Ghrelin, acylated | 1.20 | 4.84 | 0.8051 | 1.39 | 0.60 | 0.0234 | 0.31 | 0.61 | 0.6056 |
| GLP-1 | **-0.09** | **0.03** | **0.0057** | **-0.01** | **0.005** | **0.0076** | -0.001 | 0.005 | 0.7115 |
| PYY | -0.07 | 0.07 | 0.2819 | -0.01 | 0.01 | 0.1505 | -0.001 | 0.01 | 0.8899 |
|  | Posterior |  |  | Tubular inferior |  |  | Tubular superior |  |  |
| | $\beta$ | S.D. | *p-value* | $\beta$ | S.D. | *p-value* | $\beta$ | S.D. | *p-value* |
| Ghrelin, total | -0.96 | 1.66 | 0.5665 | 2.22 | 1.88 | 0.2406 | -1.11 | 1.80 | 0.5397 |
| Ghrelin, acylated | -1.99 | 1.93 | 0.3038 | 2.15 | 1.99 | 0.2827 | -0.62 | 1.82 | 0.7363 |
| GLP-1 | -0.02 | 0.01 | 0.0856 | -0.03 | 0.01 | 0.0223 | -0.02 | 0.01 | 0.1376 |
| PYY | -0.03 | 0.03 | 0.3081 | -0.02 | 0.03 | 0.4311 | -0.02 | 0.03 | 0.4003 |

$\beta$ values from the mixed-effects models for the whole hypothalamus and its subunits. Results that were significant after FDR correction ( $p < 0.05$ ) are shown in bold. Ghrelin (total and acylated) was measured after a 12h fast, while GLP-1 and PYY were measured in postprandial. S.D. : Standard deviation. GLP-1 : glucagon-like peptide 1, PYY : peptide YY.

## 4 Discussion

In this study, a tool based on a deep convolutional neural network was used to examine hypothalamic volume changes following significant weight loss induced by bariatric surgery. Our findings revealed reduced volumes of the whole left hypothalamus, specifically in the left anterior-superior and left posterior subunits. These hypothalamic morphological changes post-surgery were associated with the percentage of total weight loss, as well as with improvements in cardiometabolic variables, namely HDL-C and SBP. We also examined the associations between hypothalamic volume changes and gastrointestinal appetite-regulating hormone levels to explore potential mechanisms underlying hypothalamic alterations. We found that reduced volumes in the left anterior-superior and left posterior subunits were associated with changes in PYY levels.

To our knowledge, only one recent study has assessed hypothalamic volume following bariatric surgery using a deep convolutional neural network method (Pané et al., 2024). They found reduced volume in the whole left hypothalamus and left subunits one-year post-surgery, but no volume changes in the whole right hypothalamus, which aligns with our findings. However, they reported more widespread changes in hypothalamic volumes compared to our study. Their larger sample size one year post-surgery (56 participants), along with differences in the types of bariatric surgeries performed and the proportion of female and male participants, could potentially explain the variations in results. It is also possible that our analyses were underpowered to capture differences in volumes of certain subunits due to the smaller sample size, particularly at 24 months post-surgery.

The posterior subunit notably encompasses the lateral nuclei. The orexin neurons within these nuclei are well known for their role in food intake (Toshinai et al., 2003). Studies have reported bilateral increases in the posterior subunits in obesity (Alzaid et al., 2024; Brown et al., 2023). However, our results show that only the volume of the left posterior subunit is reduced following bariatric surgery, which is consistent with findings from a similar study (Pané et al., 2024). This suggests that alterations observed in obesity are reversible only for the left side. Lateralization of the hypothalamus has also been noted in other volumetric studies (Pané et al., 2024; Ruzok et al., 2022) and in studies assessing hypothalamic inflammation using other MRI techniques (Kreutzer et al., 2017; Schur et al., 2015; Thomas et al., 2019). Moreover, pathways involving the hypothalamus and other cerebral components participating in homeostatic feeding are suggested to be lateralized to the left side (Castro, Cole, & Berridge, 2015; Kiss et al., 2020). Thus, this lateralization could potentially explain why only the volume of the left posterior subunit decreases following bariatric surgery, as it profoundly affects the balance of food intake.

Additionally, we found a reduced volume in the left anterior-superior subunit at 4, 12 and 24 months following bariatric surgery. Similar results were reported by Pané *et al*., as they also observed reduced volume in this subunit 12 months post-surgery (Pané et al., 2024). The left anterior-superior subunit includes parts of the paraventricular nucleus, which plays a role in food intake through second order neurons that receive projections from orexinergic neurons expressing Neuropeptide Y(NPY)/ agouti-related peptide (AgRP) of the arcuate nucleus (Li et al., 2023; Roger et al., 2021). Interestingly, evidence shows a higher total volume in the anterior-superior subunits (including both left and right parts) in participants living with obesity as compared to lean controls (Alzaid et al., 2024). However, another study found no difference when examining the left and right sides independently (Brown et al., 2023). These discordant results may be explained by the small size of this subunit. Segmentation of the anterior-superior and anterior-inferior subunits is reported to have lower accuracy due to the challenging boundary between them (Billot et al., 2020).

Few studies have reported increased hypothalamic volume of the tubular inferior subunit among participants with obesity compared to those with normal BMI (Alzaid et al., 2024; Pané et al., 2024). This subunit includes, amongst other, the arcuate nucleus and the ventromedial nucleus, both known for their roles in homeostatic feeding. Our results showed no significant volume differences in this subunit following bariatric surgery, contrasting with another study that found decreased volume in individuals living with obesity but without type 2 diabetes, with no changes observed in those with type 2 diabetes (Pané et al., 2024). These differences suggest that some clinical characteristics and inter-individual variability may influence the reversibility of hypothalamic alteration in the arcuate nucleus in the short term, which could explain the lack of volume changes observed in our study. The tubular inferior subunit also includes four other nuclei, potentially explaining the absence of volume changes post-surgery in our study. The absence of recuperation of this structure could also provide insights into the mechanisms explaining frequent weight regain observed in the years following bariatric surgery. Therefore, it is important to address this gap in our current knowledge, and further studies should aim to determine whether these alterations can be reversed over a longer period.

Several mechanisms have been proposed to explain hypothalamic alterations associated with obesity. Most evidence points toward hypothalamic inflammation and gliosis, which have been demonstrated in preclinical studies and confirmed in humans using neuroimaging studies (Sewaybricker et al., 2023). Neuroinflammation is known to lead to production of proteases and free radicals that can affect the integrity of multiple brain structures. For instance, proteases could disrupt the tight junctions of the blood-brain barrier, leading to buildup of extracellular fluid and subsequent increases in volume (Rosenberg & Yang, 2007). This is consistent with the increased mean diffusivity in the hypothalamus observed in obesity, which as been reported with concomitant increased volumes (Pané et al., 2024). Moreover, alterations in perineuronal nets, an extracellular matrix protecting synapses, have been observed in rodents on a high-fat, high-sugar diet (Reichelt, Hare, Bussey, & Saksida, 2019). Without the protection of perineuronal nets, neurons are more exposed to free radicals, potentially leading to hypothalamic inflammation and increased volume. As bariatric surgery leads to decreased systemic inflammation (Askarpour, Khani, Sheikhi, Ghaedi, & Alizadeh, 2019) and reduced hypothalamic inflammation (Hankir et al., 2019; Pané et al., 2024), this reduction could explain the following decrease in hypothalamic volume post-surgery. However, our study was not designed to explore the pathways underlying these volume changes. Future studies are therefore needed to elucidate these intriguing mechanisms.

To our knowledge, our study is the first to explore the associations between hypothalamic volume changes and gastrointestinal appetite-regulating hormone levels after bariatric surgery.

We found that higher levels of PYY were associated with more pronounced reductions in the left anterior-superior and left posterior subunits post-surgery. While the physiological significance of these associations in our study is limited, it is interesting that previous studies have demonstrated the presence of PYY in the paraventricular nucleus in humans using immunocytochemistry (Morimoto et al., 2008). We also found a positive association between total ghrelin levels and volume change in the right anterior-inferior subunit, as well as a negative association between GLP-1 levels and volume change in the whole right hypothalamus and in the right anterior-inferior subunit following bariatric surgery. This direction of association is expected, as many hypothalamic subunit volumes, as well ghrelin levels are known to decrease after bariatric surgery, whereas GLP-1 levels are known to increase (Huang et al., 2023). A recent study also reported a correlation between total ghrelin levels and the volume of the anterior-inferior subunit in individuals with obesity (Alzaid et al., 2024). However, since no significant changes were observed in the subunits of the right hypothalamus following bariatric surgery, these results need to be interpreted with caution.

Several limitations need to be acknowledged. The tool we used did not allow for the segmentation of individual nuclei. Subunits include two to five nuclei, and some nuclei are divided into two different subunits, thus restricting the interpretation of the results. Our study did not include any peripheral or cerebral markers of inflammation, nor did it incorporate MRI sequences to assess brain inflammation, a potentially critical mechanism suggested to participate in the remodeling of the hypothalamus. Thus, we cannot assess the role of inflammation in hypothalamic volume changes. While this is the first study to examine long-term changes in hypothalamic structure and subregions following bariatric surgery using MRI scans up to two years post-surgery, the cohort completing the full two-year follow-up was relatively small. Nonetheless, our study is the first to assess segmentation quality and to incorporate it into our analyses.

## 5. Conclusion

In conclusion, our study shows reductions in volumes of the left hypothalamus and some specific left subunits following bariatric surgery, suggesting a recovery of hypothalamic alterations associated with obesity. Volume reductions were observed in the left anterior-superior subunit, which includes the paraventricular nuclei, and in the left posterior subunit, housing the lateral nuclei. Both of these hypothalamic nuclei play crucial roles in regulating homeostatic food intake. Our findings suggest that changes in the hypothalamus in post-bariatric surgery may be involved in shifts that could influence food intake regulation and potentially impact weight loss outcomes.

## Data Availability

All data produced in the present study are available upon reasonable request to the authors.

## Data and Code Availability

The automated hypothalamus segmentation method is publicly available at : https://github.com/BBillot/hypothalamus_seg. Data will be made available on request.

## Author contributions

Conceptualization: LB, AT, AM. Methodology: AL, JD, JF, MD, AM, MD. Data collection (participants): MP, LB. Data analysis: AL, ACC, MD, AC, JM. Writing – original draft preparation: AL, MD, AM. Writing – review and editing: AL, JD, MP, AC, ACC, LB, JM, AT. MD, AM. Supervision: AT, AM.

## Acknowledgements

We would like to recognize the contribution of surgeons, nurses and the medical team of the bariatric surgery program at IUCPQ. We would also like to acknowledge the help of MRI technicians, Xavier Morel, Coordinator of the *Plateforme d’imagerie avancée* at IUCPQ, Guillaume Gilbert, *Phillips*, Serge Simard, statistician, as well as the help of Lucie Bouffard from Dr Carpentier lab for the measurement of gut hormones and Mélanie Nadeau for the coordination of the study. Finally, we would sincerely like to thank all the participants who took part in this study.

