## Supplemental files for "Changes in hypothalamic subunits volume and their association with metabolic parameters and gastrointestinal appetite-regulating hormones following bariatric surgery"

### Supplemental material

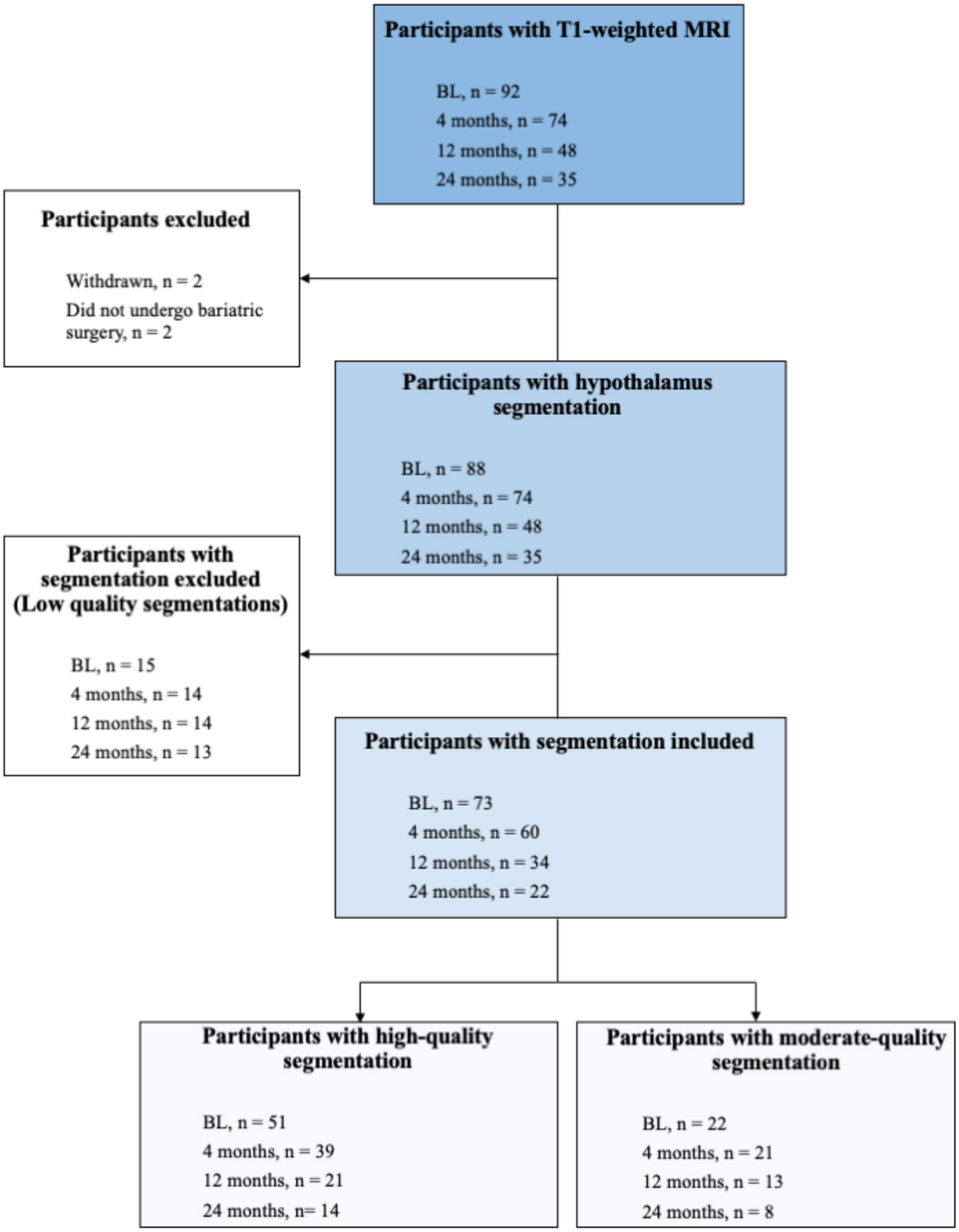


**Figure S1. Flow-chart of participants included in the study.**
BL: baseline.

**Table S1. Comparison of participant characteristics: high-quality vs. moderate-quality segmentation**

| | **Baseline** | | | **4 months** | | | **12 months** | | | **24 months** | | |
|---|---|---|---|---|---|---|---|---|---|---|---|---|
| | **High-quality** | **Moderate-quality** | ***p* value** | **High-quality** | **Moderate-quality** | ***p* value** | **High-quality** | **Moderate-quality** | ***p* value** | **High-quality** | **Moderate-quality** | ***p* value** |
| N | 51 | 22 | --- | 39 | 21 | --- | 21 | 13 | --- | 14 | 8 | --- |
| Age (years) | 45.7 ± 8.3 | 41.7 ± 10.3 | 0.0798 | 46.6 ± 7.9 | 42.6 ± 9.5 | 0.0843 | 46.7 ± 9.6 | 45.9 ± 8.6 | 0.7633 | 47.9 ± 6.7 | 48.1 ± 9.0 | 0.9942 |
| Sex (F : M) | 33:18 | 19:3 | 0.0905 | 28:11 | 15:6 | >0.999 | 16:5 | 11:2 | 0.6818 | 9:5 | 6:2 | >0.999 |
| Diabetes n (%) | 15 (29) | 4 (18) | 0.3928 | 16 (41) | 1 (5) | **0.0027** | 7 (33) | 3 (23) | 0.7041 | 3 (21) | 2 (25) | >0.999 |
| Type of surgery | | | 0.8661 | | | 0.7727 | | | 0.0717 | | | 0.3873 |
| SG | 32 | 15 | | 26 | 16 | | 12 | 8 | | 7 | 6 | |
| RYGB | 8 | 2 | | 7 | 2 | | 3 | 5 | | 3 | 2 | |
| BPD-DS | 11 | 5 | | 6 | 3 | | 6 | 0 | | 4 | 0 | |
| Weight (kg) | 120.7 ±14.5 | 122.2 ± 15.3 | 0.6862 | 93.3 ± 11.8 | 98.5 ± 11.9 | 0.1145 | 80.6 ± 12.0 | 81.1 ± 13.7 | 0.9064 | 83.2 ± 16.9 | 79.7 ± 16.6 | 0.6396 |
| BMI (kg/m$^2$) | 42.9 ± 4.0 | 45.1 ± 4.2 | **0.0307** | 34.3 ± 4.1 | 34.0 ± 3.0 | 0.8373 | 29.1 ± 3.3 | 30.4 ± 6.1 | 0.4903 | 29.4 ± 5.3 | 29.0 ± 3.7 | 0.8272 |
| Waist circumference (cm) | 129.1 ± 10.3 | 131.6 ± 10.0 | 0.3371 | 111.1 ± 11.1 | 112.1 ± 9.0 | 0.7317 | 99.0 ± 10.0 | 102.6 ± 12.7 | 0.3603 | 98.7 ± 13.5 | 100.1 ± 9.3 | 0.8057 |
| Total weight loss (%) | --- | --- | --- | 21.4 ± 4.2 | 22.0 ± 3.7 | 0.5811 | 35.1 ± 7.2 | 29.0 ± 7.2 | **0.0226** | 35.7 ± 8.2 | 30.1 ± 6.1 | 0.1124 |
| Excess weight loss (%) | --- | --- | --- | 46.5 ± 10.9 | 47.2 ± 7.9 | 0.7856 | 72.4 ± 15.1 | 66.1 ± 20.2 | 0.3014 | 73.5 ± 18.8 | 69.0 ±13.5 | 0.5627 |

Values are presented as mean ± standard deviation or n (%). F: female, M: male, SG: sleeve gastrectomy, RYGB: Roux-en-Y gastric bypass, BPD-DS: biliopancreatic derivation with duodenal switch, BMI: body mass index,

**Table S2. Results of t-tests comparing hypothalamic volumes: High-quality segmentation group versus moderate-quality segmentation group for each visit**

| | Baseline | | | 4 months | | | 12 months | | | 24 months | | |
|---|---|---|---|---|---|---|---|---|---|---|---|---|
| | **High-quality** | **Moderate-quality** | ***p* value** | **High-quality** | **Moderate-quality** | ***p* value** | **High-quality** | **Moderate-quality** | ***p* value** | **High-quality** | **Moderate-quality** | ***p* value** |
| Left hypothalamus | | | | | | | | | | | | |
| Whole left | 548.53 ± 37.29 | 545.06 ± 39.25 | 0.7205 | 542.90 ± 34.45 | 544.86 ± 43.21 | 0.8479 | 535.25 ± 45.59 | 548.26 ± 39.35 | 0.4014 | 510.04 ± 36.60 | 548.71 ± 49.04 | **0.0478** |
| Anterior-inferior | 19,50 ± 4.36 | 20.33 ± 6.78 | 0.5123 | 19.65 ± 3.81 | 17.81 ± 5.09 | 0.1196 | 17.87 ± 3.62 | 20.08 ± 4.53 | 0.1262 | 17.60 ± 4.46 | 20.57 ± 5.30 | 0.1755 |
| Anterior-superior | 29.41 ± 5.13 | 29.39 ± 5.00 | 0.9912 | 28.10 ± 4.75 | 27.99 ± 5.17 | 0.9346 | 28.98 ± 4.77 | 27.11 ± 3.97 | 0.2476 | 26.90 ± 4.77 | 28.80 ± 6.95 | 0.4569 |
| Posterior | 167.02 ± 13.39 | 171.01 ± 17.90 | 0.2958 | 163.93 ± 15.08 | 165.34 ± 18.06 | 0.7483 | 161.95 ± 14.14 | 164.61 ± 12.38 | 0.5812 | 150.31 ± 14.21 | 161.10 ± 13.67 | 0.0980 |
| Tubular inferior | 188.71 ± 16.95 | 184.68 ± 22.39 | 0.4016 | 185.89 ± 13.27 | 194.67 ± 20.57 | 0.0874 | 182.76 ± 15.94 | 192.96 ± 22.00 | 0.1269 | 176.47 ± 14.91 | 194.90 ± 16.74 | 0.0147 |
| Tubular superior | 143.90 ± 14.29 | 139.64 ± 12.20 | 0.2277 | 145.34 ± 14.77 | 139.05 ± 15.19 | 0.1250 | 143.69 ± 19.07 | 143.50 ± 14.63 | 0.9757 | 138.75 ± 12.62 | 143.34 ± 17.22 | 0.4808 |
| Right hypothalamus | | | | | | | | | | | | |
| Whole right | 540.19 ± 38.73 | 543.03 ± 37.02 | 0.7712 | 545.38 ± 39.88 | 537.37 ± 40.35 | 0.4629 | 539.79 ± 46.14 | 551.66 ± 42.55 | 0.4585 | 514.42 ± 37.88 | 546.73 ± 66.20 | 0.1577 |
| Anterior-inferior | 21.73 ± 4.41 | 22.21 ± 5.69 | 0.6958 | 21.71 ± 4.63 | 20.52 ± 5.30 | 0.4035 | 21.04 ± 5.60 | 22.33 ± 4.24 | 0.5450 | 20.12 ± 5.99 | 19.06 ± 8.20 | 0.7314 |
| Anterior-superior | 28.95 ± 4.31 | 30.24 ± 3.35 | 0.2140 | 29.31 ± 5.03 | 29.08 ± 4.86 | 0.8687 | 29.75 ± 5.93 | 29.93 ± 4.54 | 0.9269 | 28.21 ± 6.13 | 29.66 ± 6.65 | 0.6117 |
| Posterior | 155.86 ± 18.19 | 161.54 ± 17.78 | 0.2220 | 157.06 ± 18.79 | 157.31 ± 15.55 | 0.9574 | 156.94 ± 14.66 | 158.28 ± 20.66 | 0.8261 | 147.40 ± 17.47 | 164.33 ± 14.43 | 0.0311 |
| Tubular inferior | 177.02 ± 15.91 | 177.03 ± 15.72 | 0.9973 | 176.36 ± 16.19 | 177.44 ± 16.12 | 0.8060 | 175.34 ± 15.80 | 182.59 ± 16.42 | 0.2092 | 165.30 ± 14.35 | 183.08 ± 24.32 | 0.0420 |
| Tubular superior | 156.63 ± 13.66 | 152.00 ± 11.20 | 0.1669 | 160.95 ± 12.51 | 153.01 ± 15.86 | 0.0373 | 156.73 ± 19.65 | 158.53 ± 15.15 | 0.7799 | 153.39 ± 15.79 | 150.60 ± 23.15 | 0.7959 |

Values are presented as mean volume ± standard deviation. Results that were significant after FDR correction ($p<0.05$) are in bold.

**Table S3. Linear mixed-effects models showing changes in hypothalamus subunits following bariatric surgery compared to baseline, controlling for the segmentation quality variable**

| | Left | | | | | | | | |
|---|---|---|---|---|---|---|---|---|---|
| | Whole left | | | Anterior-inferior | | | Anterior-superior | | |
| | β | S.D. | *p-value* | β | S.D. | *p-value* | β | S.D. | *p-value* |
| **Intercept** | **535.72** | **53.30** | **< 0.0001** | 8.93 | 6.08 | 0.1467 | **15.59** | **6.47** | **0.0186** |
| **Age** | -0.14 | 0.52 | 0.7927 | 0.09 | 0.06 | 0.1266 | 0.07 | 0.06 | 0.2468 |
| **Sex (male)** | -3.63 | 5.10 | 0.4795 | -1.12 | 0.59 | 0.0626 | -0.66 | 0.63 | 0.3025 |
| **Baseline BMI** | -0.21 | 1.17 | 0.8610 | 0.10 | 0.13 | 0.4563 | 0.15 | 0.14 | 0.2815 |
| **Roux-en-Y** | -9.25 | 6.67 | 0.1696 | 0.35 | 0.75 | 0.6451 | -1.62 | 0.79 | 0.0439 |
| **BPD-DS** | -1.93 | 5.83 | 0.7422 | -0.17 | 0.68 | 0.8066 | 0.43 | 0.73 | 0.5534 |
| **4 months post-op** | -0.97 | 1.61 | 0.5520 | -0.27 | 0.22 | 0.2218 | **-0.50** | **0.20** | **0.0133** |
| **12 months post-op** | -4.22 | 2.16 | 0.0557 | -1.08 | 0.36 | 0.0039 | **-0.75** | **0.31** | **0.0169** |
| **24 months post-op** | **-8.73** | **2.12** | **0.0002** | -0.42 | 0.36 | 0.2437 | **-1.10** | **0.34** | **0.0036** |
| **Moderate-quality segmentation** | 6.67 | 4.28 | 0.1220 | 0.61 | 0.63 | 0.3308 | 0.26 | 0.60 | 0.6653 |
| | Posterior | | | Tubular inferior | | | Tubular superior | | |
| | β | S.D. | *p-value* | β | S.D. | *p-value* | β | S.D. | *p-value* |
| **Intercept** | **136.16** | **19.36** | **< 0.0001** | **213.41** | **24.99** | **< 0.0001** | **152.61** | **19.25** | **< 0.0001** |
| **Age** | 0.11 | 0.19 | 0.5529 | -0.55 | 0.24 | 0.0243 | 0.13 | 0.19 | 0.5123 |
| **Sex (male)** | -2.98 | 1.87 | 0.1156 | -1.75 | 2.38 | 0.4646 | 1.42 | 1.85 | 0.4469 |
| **Baseline BMI** | 0.24 | 0.42 | 0.5732 | -0.10 | 0.55 | 0.8523 | -0.46 | 0.42 | 0.2749 |
| **Roux-en-Y** | -5.26 | 2.312 | 0.0274 | 0.72 | 3.12 | 0.8189 | -6.98 | 2.40 | 0.0050 |
| **BPD-DS** | -0.05 | 2.12 | 0.9828 | -4.14 | 2.76 | 0.1373 | 0.84 | 2.11 | 0.6922 |
| **4 months post-op** | -1.32 | 0.70 | 0.0646 | 0.89 | 0.75 | 0.2438 | 0.32 | 0.75 | 0.6730 |
| **12 months post-op** | **-3.11** | **0.74** | **0.0001** | 0.80 | 0.90 | 0.3777 | 0.41 | 0.98 | 0.6748 |
| **24 months post-op** | **-6.39** | **1.25** | **< 0.0001** | -0.99 | 0.98 | 0.3195 | 0.03 | 0.74 | 0.9624 |
| **Moderate-quality segmentation** | 1.61 | 1.84 | 0.3834 | 1.71 | 2.01 | 0.3975 | 3.14 | 1.76 | 0.0784 |
| | Right | | | | | | | | |
| | Whole right | | | Anterior-inferior | | | Anterior-superior | | |
| | β | S.D. | *p-value* | β | S.D. | *p-value* | β | S.D. | *p-value* |
| **Intercept** | **545.44** | **52.45** | **< 0.0001** | 9.73 | 6.12 | 0.1166 | **20.51** | **5.95** | **0.0010** |
| **Age** | -0.74 | 0.51 | 0.1503 | -0.08 | 0.06 | 0.1722 | -0.04 | 0.06 | 0.4804 |
| **Sex (male)** | -7.39 | 5.01 | 0.1443 | -0.51 | 0.59 | 0.3841 | -0.13 | 0.56 | 0.8239 |
| **Baseline BMI** | 0.41 | 1.15 | 0.7250 | 0.31 | 0.13 | 0.0245 | 0.22 | 0.13 | 0.0949 |
| **Roux-en-Y** | -5.11 | 6.56 | 0.4389 | -0.34 | 0.76 | 0.6558 | -0.56 | 0.74 | 0.4551 |
| **BPD-DS** | -4.39 | 5.74 | 0.4461 | -1.61 | 0.67 | 0.0191 | -0.47 | 0.64 | 0.4649 |
| **4 months post-op** | 1.52 | 1.78 | 0.3973 | -0.15 | 0.25 | 0.5525 | 0.25 | 0.24 | 0.2997 |
| **12 months post-op** | 0.34 | 1.90 | 0.8582 | -0.34 | 0.33 | 0.3165 | -0.12 | 0.34 | 0.7228 |
| **24 months post-op** | -1.54 | 3.54 | 0.6668 | -0.74 | 0.39 | 0.0667 | -0.26 | 0.50 | 0.6063 |

| | | | | | | | | | |
|---|---|---|---|---|---|---|---|---|---|
| **Moderate-quality segmentation** | 3.32 | 4.54 | 0.4674 | -0.65 | 0.67 | 0.3395 | 0.49 | 0.68 | 0.4671 |
| | **Posterior** | | | **Tubular inferior** | | | **Tubular superior** | | |
| | **β** | **S.D.** | ***p-value*** | **β** | **S.D.** | ***p-value*** | **β** | **S.D.** | ***p-value*** |
| **Intercept** | **184.62** | **23.58** | **<.0001** | **164.81** | **21.92** | **< 0.0001** | **152.82** | **19.37** | **< 0.0001** |
| **Age** | -0.33 | 0.23 | 0.1620 | -0.34 | 0.21 | 0.1189 | 0.07 | 0.19 | 0.7217 |
| **Sex (male)** | -4.65 | 2.26 | 0.0436 | -1.63 | 2.10 | 0.4398 | -0.48 | 1.83 | 0.7943 |
| **Baseline BMI** | -0.35 | 0.52 | 0.4979 | 0.49 | 0.48 | 0.3131 | 0.004 | 0.42 | 0.9918 |
| **Roux-en-Y** | -1.20 | 2.93 | 0.6836 | -2.26 | 2.74 | 0.4135 | -2.01 | 2.43 | 0.4109 |
| **BPD-DS** | 0.71 | 2.59 | 0.7848 | -3.03 | 2.39 | 0.2095 | 0.55 | 2.09 | 0.7919 |
| **4 months post-op** | -0.07 | 0.66 | 0.9181 | -0.18 | 0.68 | 0.7902 | 1.33 | 0.70 | 0.0619 |
| **12 months post-op** | -0.14 | 0.76 | 0.8552 | 0.61 | 0.91 | 0.5040 | 0.65 | 1.11 | 0.5625 |
| **24 months post-op** | 0.13 | 1.30 | 0.9187 | -1.44 | 1.28 | 0.2686 | 0.62 | 1.33 | 0.6451 |
| **Moderate-quality segmentation** | -0.03 | 1.89 | 0.9879 | 1.67 | 1.97 | 0.3978 | 0.49 | 2.01 | 0.8088 |

β values from the mixed-effects models for the whole hypothalamus and its subunits. Results that were significant after FDR correction (p<0.05) are shown in bold. S.D : Standard deviation. BMI : Body mass index, BPD-DS : biliopancreatic diversion with duodenal switch.
